# Physical and sociocultural factors influencing hand hygiene behavior during COVID-19 pandemic: A cross-sectional study among traders in Lusaka District, Zambia

**DOI:** 10.1101/2024.03.06.24303900

**Authors:** Joyce Namukonda Siame, Liyali Libonda, Kanyata Kanyata

## Abstract

The COVID-19 epidemiology highlights the challenges for containment, mitigation, and control in low-resource settings like Zambia and suggests that the most vulnerable groups, particularly those living in low-income environments and depending on casual livelihoods, demand attention. This study aimed to determine factors that affect hand hygiene behavior during the COVID-19 Pandemic among traders at the New Soweto Market in Lusaka District. A cross-sectional analytical design was used to quantitatively investigate physical and sociocultural factors that affect hand hygiene behaviour during the COVID-19 pandemic among traders at New Soweto Market in Lusaka District, Zambia. The study sample included a total of 125 registered traders at the New Soweto Market, which is the biggest market in the capital city of Zambia. Majority of the participants (98.1%) demonstrated knowledge on hand hygiene. Remarkably, hand hygiene compliance was significantly associated with acquisition of hand hygiene knowledge [X (6, N = 120) = 41.49, p = 0.000] and cultural customs [X (6, N = 120) = 62.09, p = 0.000]. Further, hand hygiene compliance was significantly associated with availability of hand hygiene services or facilities within the market, [X (6, N = 120) = 13.30, p = 0.038]. Despite this, majority of the participants (61.7%) reported that they did not benefit from any hand hygiene services or facilities from the Local Authority or Ministry of Health. For an effective infection prevention and control program regarding COVID-19 pandemic, relevant authorities should consider supporting traders with hand hygiene related services.

## 1.0. Introduction

The COVID-19 epidemiology indicates that the disease is predominantly transmitted through respiratory droplets passed either directly through close, unprotected contact between an infected and a susceptible individual or indirectly when contaminated hands touch the mucosa of the mouth, nose, or eyes[1]. The Severe Acute Respiratory Syndrome-associated coronavirus (SARS-CoV-2), being the virus responsible for COVID-19 disease, can spread from contaminated hands to surfaces, further facilitating indirect transmission[2]. Although it may seem that a person is most contagious when they are sick, there is some evidence that suggests transmission can happen before symptoms appear[3]. In addition, the virus is detectable for some time on surfaces, aerosols, and stool [4].

Along with social distancing, hand hygiene has been repeatedly advised as one of the key actions to reduce transmission of the SARS-CoV-2 virus, responsible for the COVID-19 pandemic[5]. Practicing hand hygiene, which includes the use of alcohol-based hand sanitizer (ABHS) or hand washing with soap and water, non-shaking of hands, and avoidance of coughing directly on hands, is a simple and yet effective way to prevent the spread of COVID-19 in high-risk settings [6]. Therefore, frequent, and proper hand hygiene behavior is considered one of the most important control measures for preventing infection with COVID-19, including in community settings [7].

Nevertheless, hand hygiene may not always be taken as seriously as it should, with compliance and adherence in clinical and public settings far from optimal over time [8,9]. Multiple reports from different countries have shown that the hand hygiene compliance rate has been estimated at only 40% [10], while the rate of adherence in critical care units was only 46.25% [8]. Although this is a simple and lifesaving task, it is not, regrettably, always undertaken[11]. Despite stakeholders endeavoring to ensure the focus on hand hygiene continues, the transmission dynamics of the disease may be slowly influencing lower risk perceptions and reluctance to observe such an important measure [12]. This emphasis on tackling communicable disease prevention might not differ from what is typical in Zambia.

Furthermore, Lusaka’s New Soweto Market, located west of the Central Business District is one of the most frequently populated trading places in Lusaka. However, hygiene conditions continue to be in a dire state. The trading spaces are usually overcrowded, with traders and consumers more likely to be in contact with each other through the exchange of cash notes, handshakes, and touching of contaminated surfaces. Basic hand sanitation amenities, including hand hygiene facilities, appear to be in short supply at the market and likely to negatively influence hand hygiene behaviour. Bezerra et al. [8] and Pittet [9] assert that hand hygiene may not always be taken as seriously as it should be in terms of compliance and adherence in public settings such as markets. In a situation observed in different countries, space constraints and overcrowding lead to direct and indirect human contact in markets and dwellings, making the spread of an infection highly likely [13]. As such, Soweto Market presents an unpleasant trading environment with the potential of exacerbating person-to-person transmission of COVID-19 in Lusaka District.

This study aimed at determining physical and sociocultural factors that affect hand hygiene behaviour during the COVID-19 pandemic among traders at New Soweto Market. This was achieved by investigating the influence of social demographic characteristics, hand hygiene knowledge, sociocultural factors, and effects of the physical environment on the hand hygiene behaviour of the traders.

### 1.1. Aim of the study

To investigate physical and sociocultural factors that affect hand hygiene behaviour during the COVID-19 pandemic among traders at New Soweto Market in Lusaka District.

## 2.0. Materials and Methods

### 2.1. Study design

A cross-sectional analytical design was employed to investigate the extent to which the variables of social demographic, sociocultural, and physical environmental factors influence hand hygiene behavior among traders at the New Soweto Market during the COVID-19 pandemic.

### 2.2. Study area and period

The study was conducted at the new Soweto Market in Lusaka District. Recruitment of participants and data collection took place between January 3, 2022, and January 31, 2022. With a population of more than 2.5 million, Lusaka is both the capital and biggest city of Zambia [14,15]. Zambia’s largest formal, council-managed, open-air urban market, Soweto Market, is situated close to Lusaka’s Central Business District (CBD). It serves as both a wholesale and retail space for a variety of food items farmed and produced within and outside Lusaka [16]. The Market plays a significant role in ensuring food security in Lusaka, as it serves as the main landing point for food coming from farmers and dealers in the greater Lusaka City area and beyond [14]. A wide range of players are interested in the Soweto Market’s operations because of its central position in shaping urban food systems and the clear money flows in Lusaka’s food value chain, which raise the stakes[13].

### 2.3. Inclusion and exclusion criteria

This study was designed to include all registered traders who were trading from the New Soweto market in Lusaka District during the COVID-19 pandemic and who were older than 18 years. Individuals who were physically or mentally ill and younger than 18 were excluded from participation. Furthermore, since participation in the study was entirely voluntary, individuals who refused to provide their consent were not allowed to participate.

### 2.4. Sample size determination

A sample size of 125 was chosen from a table of estimated adequate sample sizes for cross-sectional studies with a fixed value of significance level (α = 0.05), standard deviation (2.5), and margin of error considered as 10% of the variance [17]. However, five female participants withdrew during the data collection process, bringing the total number of participants to 120. To avoid gender bias, this sample was stratified in proportion to the general market population and consisted of 60 male and 65 female traders. Consideration for the sample size chosen was also made in view of limited time and financial resources.

### 2.5. Sampling Procedure

Participants in this study were chosen using a proportionate stratified sampling approach according to their gender. This procedure was chosen because it provided each element in the population an equal chance to be selected as a study participant. The sampling procedure involved dividing the entire population of registered traders into homogeneous groups (strata) based on gender, this was followed by taking random samples from the two stratified groups, in proportion to the population of both male and female registered traders at the market.

### 2.6. Data collection tools and procedures

Upon gaining access to the market facility, with permission from the Lusaka City council, those who met the inclusion criteria were then approached with the help of market officials and asked if they were willing to participate. An information sheet explaining the nature and purpose of the study was given to the participants after which, those who volunteered to participate in the study had to provide written consent.

A structured questionnaire and an observation checklist were used to collect data. The questionnaire items were adopted from a WHO longitudinal survey on monitoring behavioural insights for use during the COVID-19 pandemic [9]. A subset of the WHO questions was used to allow focus on the variables relevant to this study. Adaption of the instrument was conducted to ensure consistency with the Zambian context, with a focus on social demographic, hand hygiene information, social cultural and physical environmental factors that may affect hand hygiene behavior among traders at Soweto Market during the COVID-19 pandemic. Items for the checklist were developed consistently with the Ministry of Health standards and guidelines for infection prevention and control[14].

A pilot study was conducted at the old Soweto Market in Lusaka District over a two day period, from 13^th^ to 14^th^ December, 2021. This is a different site from the actual study site to avoid getting preconceived responses. Ten percent (12 participants) of the sample size for the study was selected to participate in the pilot study. Scale reliability of the questionnaire was measured using SPSS with a 0.80 Cronbach alpha value.

### 2.7. Data quality control

The WHO-adapted questionnaire on monitoring hand hygiene behavioural insights was pre-tested among 12 participants at a similar busy market in Lusaka district. The two data collectors for this study were trained over a three-day period before the pilot study was conducted by the principal investigator. A formative data analysis was then conducted to ascertain the validity and reliability of the questionnaire. The principal investigator conducted onsite spot-checking for incomplete questionnaires to ensure correct data entry. The supervisors controlled the overall data collection process.

### 2.8. Data analysis

The chi-square test was selected as an appropriate statistical method for this study because it met the following assumptions: random selection of study participants; both the dependent and independent variables were categorical data. For the analysis, the data was first entered and cleaned in Microsoft Excel. It was then exported into the Statistical Package for Social Sciences (SPSS) version 26, which was utilised for descriptive and inferential statistical analysis. A comprehensive characterisation and summarisation of the socio-demographic data was conducted by calculating frequencies and percentages. The Chi-square test was used to evaluate the statistical significance of the association between the dependent variable (hand hygiene behaviour) and independent variables (social demographic characteristics, hygiene knowledge, sociocultural factors and the physical environment).

## 3.0 Ethics approval and consent to participate

Ethical approval was obtained from the University of Zambia Zambia Natural and Applied Sciences Research Ethics Committee. Clearance was also sought from the Lusaka City Council and leadership at New Soweto Market. In addition, permission was obtained from each individual participant. Written informed consents were also obtained from all the participants. Participants were informed of their voluntary participation and that participation was without any material or financial benefits. However, it was also explained that the study was expected to have long-term benefits in terms of informing innovative approaches to hand hygiene as well as a policy shift during pandemics such as COVID-19. Confidentiality regarding information to be shared by participants was maintained throughout the study. No names of participants were recorded anywhere for the purpose of confidentiality. The participants filled in the questionnaire in their free time within the shortest possible time. Further, they were not subjected to any physical harm as the study did not involve any invasive procedures.

## 4.0 Results

### 4.1. Socio-demographic characteristics of respondents

The targeted number of participants was 125, but five withdrew for various personal reasons such as having very busy schedules at the market and attending to family problems, leaving a total of 120 participants. The majority (71.7%) of the participants were aged 25 years and above. Regarding level of education, 61.7% had attained secondary school education and were mostly married (49%). Furthermore, a bigger percentage (93.3%) of the participants belonged to the Christian faith group, while a small section were Muslims, Hindus, and other respective faith groups (Table.1).

**Table 1.** Participants’ socio-demographic characteristics (N=120)

| Variable | N | (%) |
| --- | --- | --- |
| **Age** |  |  |
| Below 24 yrs. | 34 | 28.3 |
| 25 yrs. and above | 86 | 71.7 |
| **Gender** |  |  |
| Male | 60 | 50 |
| Female | 60 | 50 |
| **Education Level** |  |  |
| Primary | 18 | 15 |
| Secondary | 74 | 61.7 |
| College | 17 | 14.2 |
| University | 11 | 9.2 |
| **Marital Status** |  |  |
| Single | 49 | 40.8 |
| Married | 60 | 50 |
| Widowed/Divorced | 11 | 9.2 |
| **Religious Affiliation** |  |  |
| Christian | 112 | 93.3 |
| Muslim | 6 | 5.0 |
| Hindu | 1 | .8 |
| Others | 1 | .8 |

### 4.2. Socio-cultural factors, knowledge, and physical environment related to hand hygiene behaviour

The majority (93.3%) of participants who had complied with hand hygiene were Christians as compared to other religions. Chi-square test revealed a significant association between hand hygiene compliance and religious affiliation, [X (9, N = 120) = 20.02, p = 0.018]. Further, the evidence indicates that most participants (92.5%) who complied with hand hygiene practice are those who believed that religion could improve hand hygiene practice as compared to those who did not (8.1%). Compliance with hand hygiene practice was significantly associated with participants’ belief that religion can improve hand hygiene practice, [X (9, N = 120) = 57.59, p = 0.000].

An analysis of the effect of knowledge on hand hygiene practice indicated that the majority (91%) of participants had acquired information on hand hygiene and only (9.2%) did not acquire this information. The association between hand hygiene compliance and acquisition of hand hygiene knowledge was statistically significant, [X (6, N = 120) = 41.49, p =.000]. It is also interesting that 78.3% of the participants who complied with hand hygiene also reported cultural customs not to conflict with the prevention of COVID-19, while 21.7% stated cultural customs to be in conflict. Compliance with hand hygiene practice was significantly associated with existence of conflicting customs, [X (6, N = 120) = 62.09, p = 0.000].

The impact of availability of hand hygiene services and practice was evaluated. The evidence indicated that 72.5% of the participants reported to have not received any hand hygiene services in relation to hand hygiene behaviour, while a small number (27.5%) reported having received such services. The Chi-square test revealed a significant association between hand hygiene compliance and availability of hand hygiene services [X (6, N = 120) = 13.30, p = 0.038]. These findings are summarized in (Table.2).

**Table 2.**
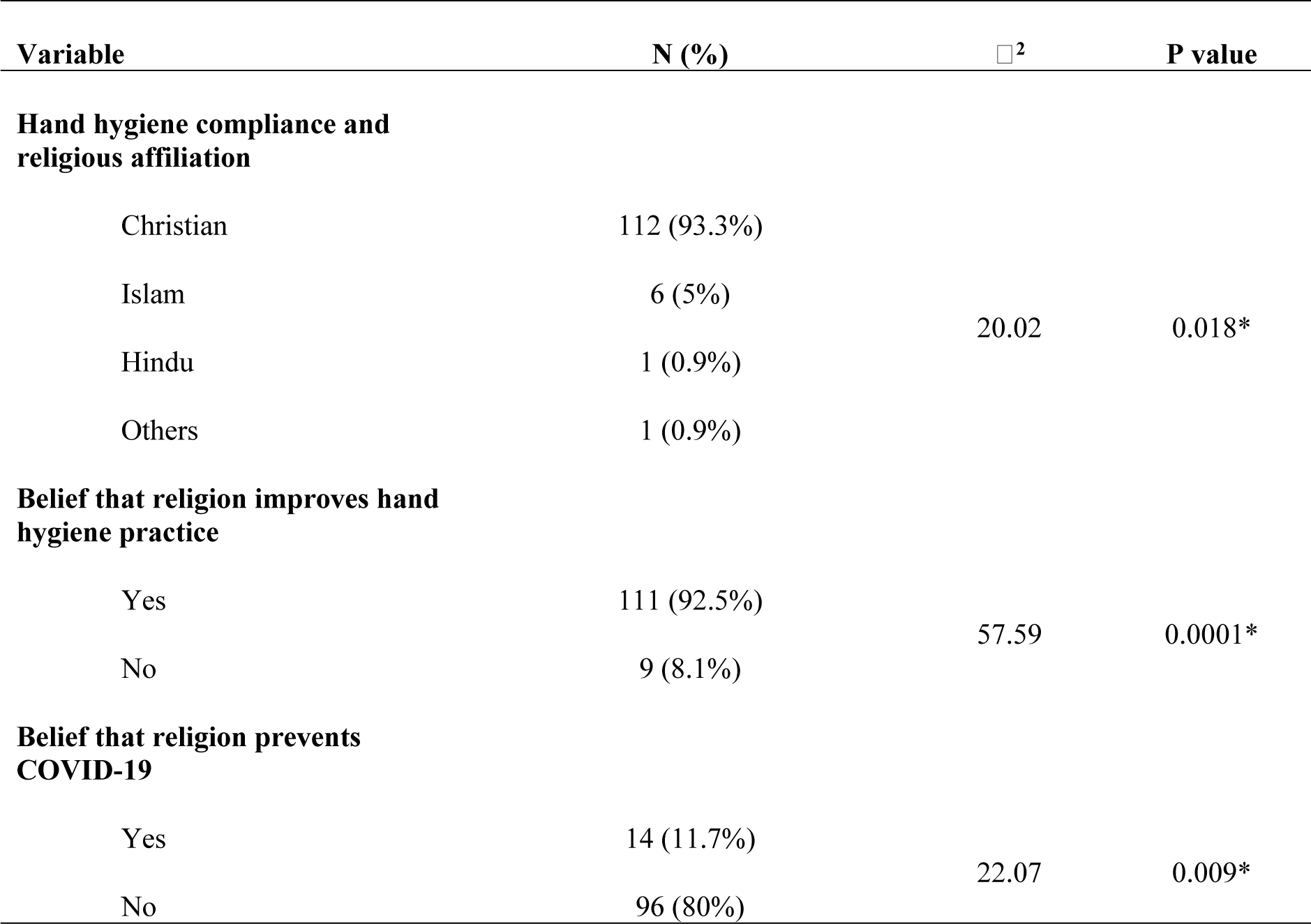
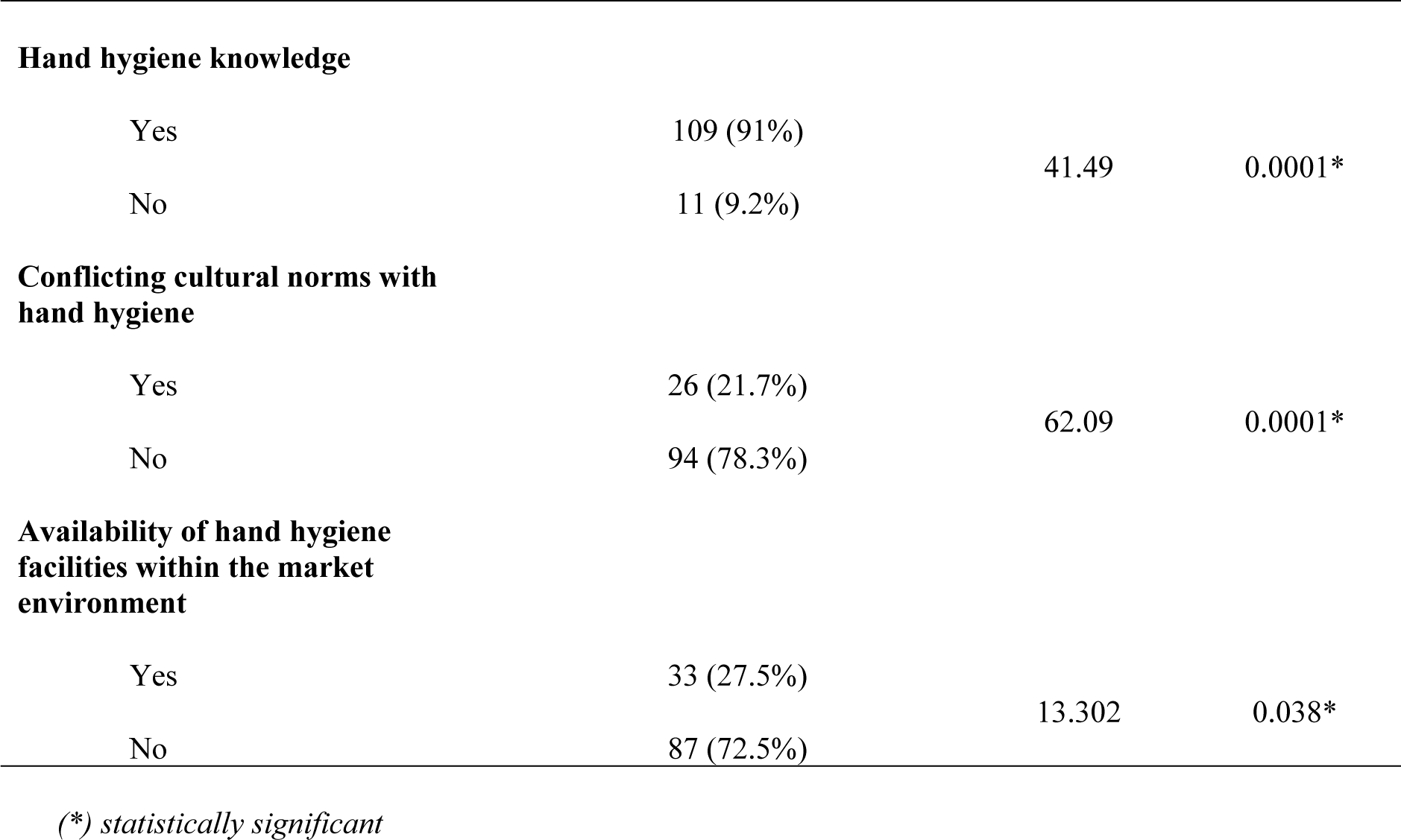
Socio-cultural factors, knowledge, and physical environment related to hand hygiene behaviour.

| Variable | N (%) | $\chi^2$ | P value |
| --- | --- | --- | --- |
| **Hand hygiene compliance and religious affiliation** |  |  |  |
| Christian | 112 (93.3%) | 20.02 | 0.018* |
| Islam | 6 (5%) |  |  |
| Hindu | 1 (0.9%) |  |  |
| Others | 1 (0.9%) |  |  |
| **Belief that religion improves hand hygiene practice** |  |  |  |
| Yes | 111 (92.5%) | 57.59 | 0.0001* |
| No | 9 (8.1%) |  |  |
| **Belief that religion prevents COVID-19** |  |  |  |
| Yes | 14 (11.7%) | 22.07 | 0.009* |
| No | 96 (80%) |  |  |
| **Hand hygiene knowledge** |  |  |  |
| Yes | 109 (91%) | 41.49 | 0.0001* |
| No | 11 (9.2%) |  |  |
| **Conflicting cultural norms with hand hygiene** |  |  |  |
| Yes | 26 (21.7%) | 62.09 | 0.0001* |
| No | 94 (78.3%) |  |  |
| **Availability of hand hygiene facilities within the market environment** |  |  |  |
| Yes | 33 (27.5%) | 13.302 | 0.038* |
| No | 87 (72.5%) |  |  |

(\*) statistically significant

## 4.0. Discussion

### 4.1. Socio-demographic characteristics

This study involved 120 participants, of which 50% were males and 50% were females. Five participants from the study sample size of 125 withdrew their participation in this study because of varying personal engagements at the time of data collection. Nevertheless, the study revealed that the majority (71.7%) of participants were aged 25 years and above, but participants’ age was not statistically significantly associated with hand hygiene behaviour. A higher proportion of participants in this majority age range is expected considering that most of the traders at the market are more likely to be adults trying to earn a living through trade at the marketplace. In addition, the study revealed that most of the participants (61.7%) had at least attained a secondary school education.

However, the association between hand hygiene behaviour and education level was not statistically significant (p>0.05). This finding, therefore, implies that traders’ education attainment may not have had an impact on their hand washing behaviour during the COVID-19 pandemic. In contrast, previous research has found a statistically significant link between age, education level, and hand hygiene behaviours, with younger people being more likely to make beneficial changes[15]. Furthermore, this study has revealed that in comparison to other religious groups, the Christian faith group had a larger proportion (93.3%) of traders who belonged to it. Taking into consideration that Zambia is predominantly a Christian nation, this outcome was not unexpected.

The study also revealed that hand hygiene compliance was significantly associated with religious beliefs (p= 0.000). In addition, the study has shown that most participants (91.8%) believed that religion can improve hand hygiene practice. This finding is in agreement with that of Stavrova [19], who through an analysis that linked religion to social pressures indicated that individuals are more likely to be committed to their values if they believed in those values at a personal level. It can, therefore, be supposed that traders’ hand hygiene behaviour at Soweto Market was more likely to have been influenced by closely held Christian values, considering that the majority (Table.1) of them belonged to this faith category. The role played by religion in influencing hand hygiene practice may also help in designing appropriate hand hygiene programs targeting specific religious groups at Soweto Market. Similarly, the present study has revealed that the majority (88.3%) of traders who observed hand hygiene behaviour also believed that COVID-19 can be prevented by observing both preventive guidelines and praying. In this regard, it can be suggested that religion was less likely to negatively influence participants’ beliefs about hand hygiene as a COVID-19 preventive measure.

### 4.2. Hand hygiene knowledge

Results of this study have revealed a statistically significant relationship between acquisition of hand hygiene knowledge and self-reported hand hygiene compliance (p = 0.000). The study also showed that the majority (98.1%) of traders at Soweto Market had acquired knowledge on hand hygiene in comparison to those who did not (7.8%). In the current study, the revelation that more traders had good knowledge levels on hand hygiene was perhaps due to their usual understanding of personal and hand hygiene which may have been obtained from formal and informal learning processes. This finding could be generally considered as having a positive influence on traders’ hand hygiene behaviour at Soweto Market during the COVID-19 pandemic.

The finding of the present study indicating a statistically significant relationship between acquisition of hand hygiene knowledge and self-reported hand hygiene compliance is consistent with that of Tan et al. [20], who found a positive relationship between hand hygiene knowledge and self-reported hand hygiene practice. Similarly, findings of a study done in Ethiopia revealed that 82.9% of participants had good knowledge and 17.1% had poor knowledge of hand hygiene [19]. Further, a Nigerian study on knowledge and hand hygiene practice among health workers revealed that 82.4% of participants had good knowledge of hand washing and 17.6% had poor knowledge. In the Nigerian study, observations on the practice of hand washing further revealed that 42.2% of participants always practised hand washing and 34.3% practiced occasionally, while 23.5% never practised hand washing [21].

### 4.3. Social-cultural factors and hand hygiene

This study has demonstrated that compliance with hand hygiene practice was significantly associated with existing cultural beliefs and customs (p=0.000). In addition, the study has shown that the majority (80.5%) of traders who complied with hand hygiene also reported that cultural customs were not in conflict with the prevention of COVID-19. Based on the findings, it can be suggested that hand hygiene behaviour among traders at Soweto Market was less likely to have been influenced by conflicting cultural beliefs and customs. Likewise, the findings also highlight the important role that cultural beliefs and customs could have played in the prevention of COVID-19 by consistently influencing good hand hygiene behaviour.

The finding in the current study that cultural beliefs and customs were significantly associated with hand hygiene practice among traders at Soweto Market is congruent with that of Davis [16], who reported that traders’ native culture influenced their hand-hygiene practice and believed that their native culture had influenced their hand-hygiene practice from early childhood. This is in line with Zhao et al. [17] findings that workers were more likely to follow hand-hygiene instructions if the intervention was linked to their own personal hygiene habits, such as past experiences, peer observation, ease of access to resources, and a need to practice hand-hygiene. This finding could mean that interventions aimed at promoting improved hand hygiene should consider the way of life of market traders to capitalize on the favourable cultural features that can aid in the prevention of COVID-19.

### 4.4. Physical environment and Hand hygiene

This study has revealed that the majority (61.7%) of the traders at Soweto Market reported that they were not provided with hand hygiene facilities or services by the Ministry of Health or Local Authority. Moreover, the study demonstrated that there was a statistically significant association between hand hygiene compliance and the availability of hand hygiene facilities or services within the market environment (p=0.038). Nevertheless, this finding in the current study is contrary to the recommendations of ILO on the necessary prerequisites for the prevention of COVID-19 in the workplace. Moreover, the finding of this study may suggest a heightened negative influence on hand hygiene by no availability of such services among traders at Soweto Market during the COVID-19 pandemic.

In terms of the standards required to maintain a conducive environment that supports hand hygiene at workplaces, ILO [24] guides that hand hygiene facilities and services need to be available in sufficient quantities at the workplace and accessible to all as part of the measures of prevention and control against COVID-19. Furthermore, ILO [24] clarifies that components that are required for such services include access to hand sanitizers, washbasins, soap, running potable water and single-use towels or other means of hand drying. It is, therefore, worth noting that workplace environments such as marketplaces can benefit more in terms of hand hygiene by ensuring that recommendations such as those of the ILO are observed during the COVID-19 pandemic.

## 5.0 Conclusion

The study suggests that Christianity, the religious faith of most traders at Soweto Market, significantly influenced hand hygiene behaviour, potentially positively influencing it during the COVID-19 pandemic. Additionally, their knowledge on hand hygiene may have influenced their behaviour. Furthermore, the results of this study led to the conclusion that cultural customs of traders did not conflict with promoting good hand hygiene behaviour, but challenges arose due to the lack of facilities or services in the physical environment of the market, which could have made it difficult for traders to prevent COVID-19 spread.

Though conclusions may be drawn from interaction of some variables in this study, the limited scope of this study in terms of non-inclusion of other similar trading spaces in the country arguably may restrict the generalisability of the study findings to other populations countrywide. Nevertheless, the researchers believe that the study has highlighted important factors that have the potential to influence hand hygiene behaviour during pandemics, such as COVID-19, in highly populated spots such as markets.

The study, though limited in scope, highlights significant factors that may influence hand hygiene behaviour during pandemics in highly populated environments. The findings could benefit future studies targeting overcrowded populations outside clinical settings, as available evidence suggests that previous studies were largely conducted in clinical contexts.

## Data Availability

Dataset for this study is available and ready to be summited upon request by the editorial team.

## 6.0 Supporting information

Supporting documents for this study include Ethics clearance letter, participants information sheet and consent form.

## 7.0 Acknowledgements

The authors express their gratitude to the Lusaka City Council staff members at the New Soweto market for the support rendered in facilitating easy access to the traders.

## 8.0 Author contributions

**Conceptualization**: Joyce Namukonda Siame.

**Data curation**: Joyce Namukonda Siame, Kanyata Kanyata and Liyali Libonda.

**Formal analysis**: Joyce Namukonda Siame, Kanyata Kanyata and Liyali Libonda.

**Methodology**: Joyce Namukonda Siame.

**Supervision**: Kanyata Kanyata and Liyali Libonda.

**Validation**: Kanyata Kanyata and Liyali Libonda.

**Visualization**: Joyce Namukonda Siame, Kanyata Kanyata and Liyali Libonda.

**Writing** – original draft: Joyce Namukonda Siame.

**Writing** – review & editing: Kanyata Kanyata and Liyali Libonda.

